# Lower Odor Identification in Subjective Cognitive Decline: A Meta-analysis

**DOI:** 10.1101/2025.04.15.25325887

**Authors:** Benoît Jobin, Coline Zigrand, Johannes Frasnelli, Benjamin Boller, Mark W Albers

## Abstract

**Introduction:** Odor identification correlates with Alzheimer’s disease (AD) biomarkers, and its decline may emerge before measurable cognitive deficits—as early as the subjective cognitive decline (SCD) stage. We aimed to compare odor identification between SCD and cognitively normal (CN) stages and investigate whether cognitive differences moderate olfactory deficits.

**Methods:** A systematic search of four databases identified studies assessing olfactory identification and cognitive screening in individuals aged 50+. A random-effects meta-analysis was performed on 11 studies (660 SCD, 574 CN).

**Results:** Individuals with SCD exhibited lower olfactory identification scores compared to CN participants (SMD = −0.67, 95%CI [−1.31, −0.03], *p* = .04). Meta-regression revealed a negative association (β = −1.79, p = .02) between cognitive and olfactory differences, indicating that greater cognitive decline was not consistently associated with greater olfactory deficits, lower odor identification scores in SCD occurred despite minimal cognitive differences across groups.

**Discussion:** Odor identification is lower in pre-MCI individuals reporting SCD. Olfactory decline may emerge independently prior to measurable cognitive decline, supporting the role of odor identification as a screen for AD.

## 1. Background

Alzheimer’s disease (AD) is a progressive neurodegenerative disease characterized by the accumulation of amyloid-β plaques and tau neurofibrillary tangles, followed by widespread neurodegeneration and cognitive decline, culminating in dementia.^1^ Early identification of individuals at risk for AD is critical for timely intervention, particularly as the disease progresses over decades before the occurrence of first cognitive deficits,^2^ and as therapies targeting amyloid pathology and other early pathological stages become available. Subjective cognitive decline (SCD) refers to self-reported cognitive decline in the absence of measurable deficits on neuropsychological tests.^3^ SCD is considered the first symptom of AD, appearing at stage 2 of the disease,^1,4^ and has been associated with an increased risk for the development of mild cognitive impairment (MCI) and progression to the dementia stages of the disease.^5,6^

While SCD is associated with an increased risk of cognitive decline, not all individuals reporting SCD will decline.^6^ Furthermore, the presence of AD biomarkers among individuals with SCD varies considerably, ranging from 10% to 76%.^5,7,8^ This broad variability suggests a lack of diagnostic precision and highlights the heterogeneity of SCD as a clinical entity. Therefore, SCD alone is not sufficient to predict cognitive decline, necessitating the identification of additional reliable, and easily accessible markers that can improve the specificity of early risk assessment.

One promising candidate for preclinical AD detection is odor identification impairment. Multiple studies have demonstrated that odor identification is impaired at the dementia^9^ and MCI^10^ stages and is thought to emerge even before measurable episodic memory deficits.^11^ The primary olfactory cortex morphometry is damaged in MCI^12^ and olfactory identification is associated with hippocampal volume and entorhinal cortex thickness in patients with MCI^13–15^ and individuals with SCD.^16^ Olfactory identification is also associated with longitudinal tau pathology accumulation within the central olfactory system in cognitively normal individuals.^17^ Several longitudinal studies have demonstrated that olfactory identification scores are associated with cognitive decline in cognitively normal individuals^18–20^ and conversion from MCI to dementia stage.^21,22^ Taken together, these findings suggest that olfactory impairment may represent a subtle, early marker of neurodegeneration, and could have predictive value in the detection of preclinical AD.

A prior meta-analysis^23^ suggested a trend toward lower olfactory identification scores in individuals with SCD compared to cognitively normal (CN) older adults. The number of included studies was small (k = 5), limiting the analysis of potential moderators and the interpretation of the conclusions. Since its publication, several new studies have examined olfactory function in SCD. With the advent of treatments that mitigate the burden of amyloid pathology at the MCI and early dementia stages, it is crucial to identify low-cost, non-invasive, and accessible signs and markers for AD that manifest prior to the onset of measurable cognitive deficits, particularly before the progression to MCI. Olfactory testing may thus offer a valuable complement to existing cognitive screening tools and could aid in stratifying risk in clinically normal individuals reporting cognitive concerns. The aim of the meta-analysis was (1) to compare odor identification between older adults with SCD and CN older adults without cognitive complaint, and (2) to evaluate the magnitude of olfactory identification impairment in SCD by comparing the effect sizes of olfactory differences with those observed in general cognitive screening measures (e.g., Montreal Cognitive Assessment - MoCA,^24^ Mini-Mental State Examination - MMSE^25^).

## 2. Methods

The protocol of this meta-analysis was not registered.

### 2.1 Study Eligibility Criteria

Eligible studies included a direct comparison of odor identification scores between individuals with SCD and CN participants without cognitive impairment or cognitive complaint, with both groups including men and women aged 50 years and older. The definition of SCD was based on two core criteria: (1) self-reported cognitive decline relative to a previous performance level and (2) normal cognitive performance on standardized cognitive tests, such as the MMSE or the MoCA to exclude MCI. CN participants were required to be CN without objective cognitive impairment (i.e., scoring > −1.5 standard deviations on cognitive tests) and without self-reported complaints of cognitive decline. Studies were excluded if participants were younger than 50 years of age, had a diagnosed cognitive impairment, or had a psychiatric or neurological condition that could affect cognitive or olfactory function.

#### Outcome

Eligible studies had to assess olfactory identification using a validated olfactory identification test to be included in the meta-analysis. The eligible tests included the Sniffin’ Sticks Identification Test (SSIT),^26^ the Odor Perception Identification Test (OPID)^13^/ the AROMHA Brain Health Test,^27^ the University of Pennsylvania Smell Identification Test (UPSIT),^28^ or the Cross-Cultural Smell Identification Test (CC-SIT).^29^ Similarly, eligible studies were required to include a measure of general cognition functioning using a standard screening test (MoCA or MMSE)^24,25^ to allow for effect size comparisons across cognitive and olfactory measures.

### 2.2 Search Strategy and Information Source

A systematic literature search was conducted to identify relevant studies published up to February 1^st,^ 2025. The search was conducted in the PsycNET, PubMed, Academic Search Complete (EBSCO), and Scopus databases. The following keywords and Boolean operators were used: (“SCD” OR “subjective cognitive decline” OR “subjective cognitive impairment” OR “subjective memory impairment” OR “subjective memory decline” OR “cognitive complaint*” OR “memory complaint*” OR “cognitive concerns” OR “memory concerns”) AND (”olfac*” OR “smell” OR “olfactory dysfunction” OR “olfactory impairment”). Studies had to be published in English.

After the removal of 106 duplicates, 113 titles and abstracts were screened for eligibility. We excluded studies that were unrelated to the research question, review articles, case studies, qualitative studies, or the absence of a control group. After the initial screening, 40 studies were identified as potentially eligible and were further assessed for inclusion. After reviewing full-text articles, 11 studies and 13 reports met the inclusion criteria (Figure 1). The list of included studies is reported in Supplementary material.

**Figure 1.**
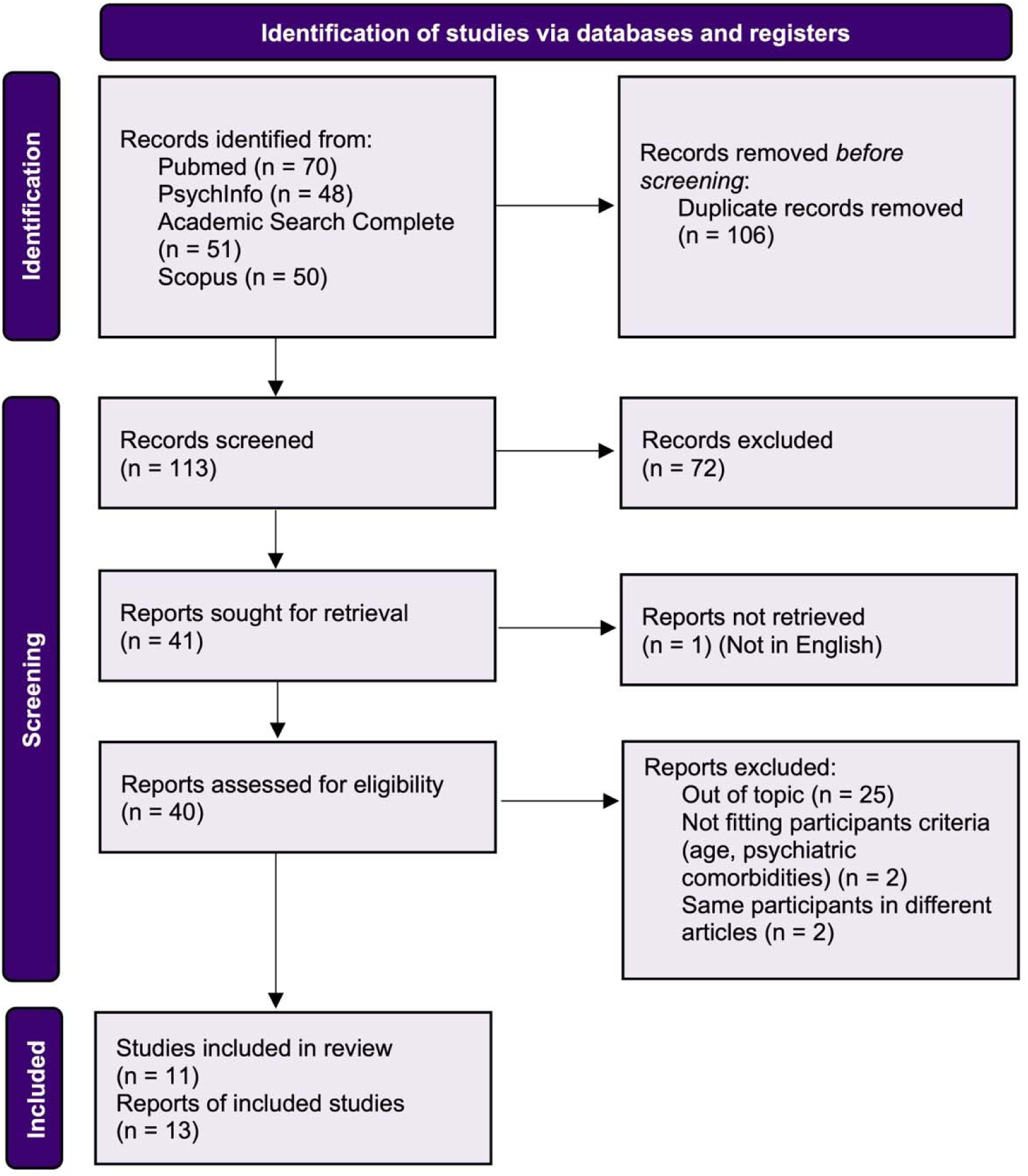
PRISMA Flow Diagram of Study Selection.

The figure illustrates the study selection process based on PRISMA (Preferred Reporting Items for Systematic Reviews and Meta-Analyses) guidelines.

### 2.3 Risk of Bias Assessment

The methodological quality of each included study was assessed using the Newcastle-Ottawa Scale (NOS), as recommended.^30^ The NOS^31^ is a tool designed to evaluate the quality of non-randomized case-control studies in meta-analyses, assessing three key domains: participant selection, comparability between groups, and outcome ascertainment. The scoring system ranges from 0 to 9 points, with higher scores indicating higher study quality. Studies scoring 0–3 were considered at high risk of bias, 4–6 at moderate risk, and 7–9 at low risk. The risk of bias for each study was independently evaluated by two reviewers (BJ and CZ). If disagreement emerged, the most conservative score was assigned. No major disagreements occurred during this process, and no studies were excluded based on quality assessment. The final risk of bias scores for each included study are summarized in Supplementary Table 1.

### 2.4. Statistical Analyses

We conducted random-effects meta-analyses using the *meta* package in R. We standardized mean differences (SMD, Hedges’ *g*) for each study by comparing the mean scores of SCD and CN groups. To improve variance estimation, we performed random-effects model using the Hartung-Knapp adjustment, with effect sizes weighted by the inverse variance of their standard errors. This approach provides robust confidence intervals and improves statistical inference, especially in meta-analyses with small numbers of studies or high heterogeneity.^32–34^ We assessed heterogeneity using Cochran’s *Q* statistic, Higgins’ *I*² index, and τ² to quantify between-study variability. High heterogeneity (*I*² > 75%) prompted further investigation through meta-regression. We conducted meta-regression analyses to examine the potential moderating effects of *age*, *sex proportion* (female), *educational level* (years), and *test modality* on effect sizes.

We assessed potential publication bias using visual inspection of funnel plots, Egger’s regression test, and the trim-and-fill method. Funnel plot asymmetry and Egger’s test *p*-values < .05 were considered indicative of potential bias. We used the trim-and-fill method to impute missing studies and adjust the pooled effect size under the assumption of publication bias.

Statistical significance was set at *p* < .05 for all analyses.

## 3. Results

### 3.1 Study selection and characteristics

A total of 11 studies met the inclusion criteria, combining 660 individuals with SCD and 574 CN participants (see Figure 1 & Table 1). Two reports/articles used the same sample, therefore this dataset counted as a single study to avoid duplication.^35,36^ According to the study quality assessment, no studies were excluded for a high risk of bias (i.e., NOS scale ≤3) (Supplementary Table 1).

**Table 1.**
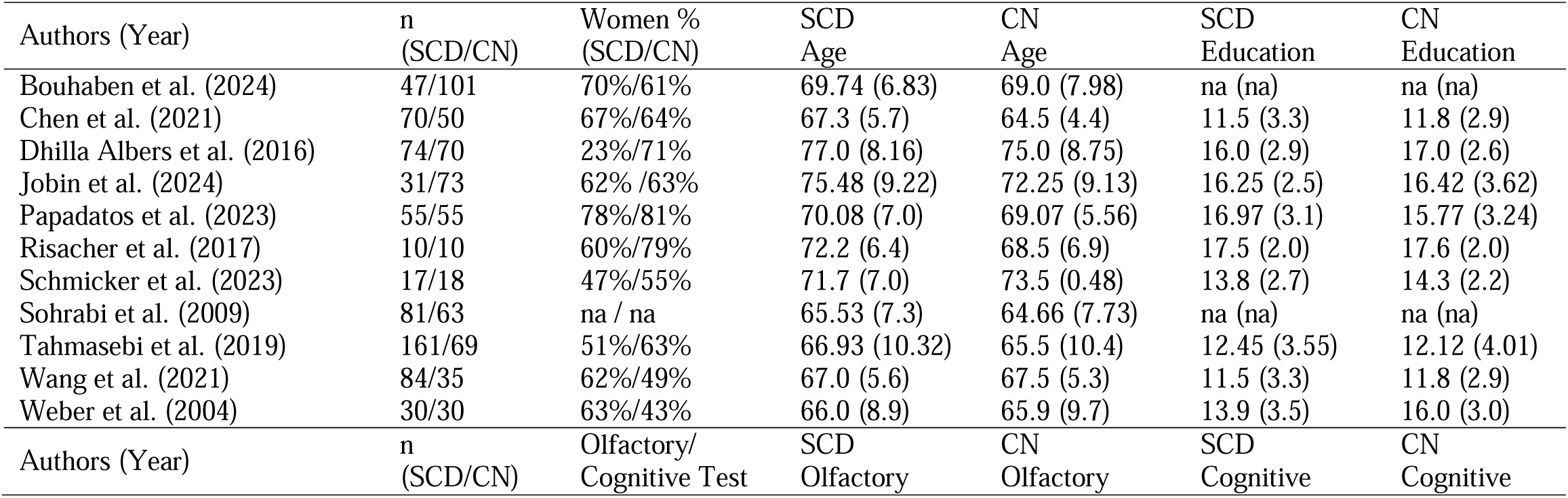
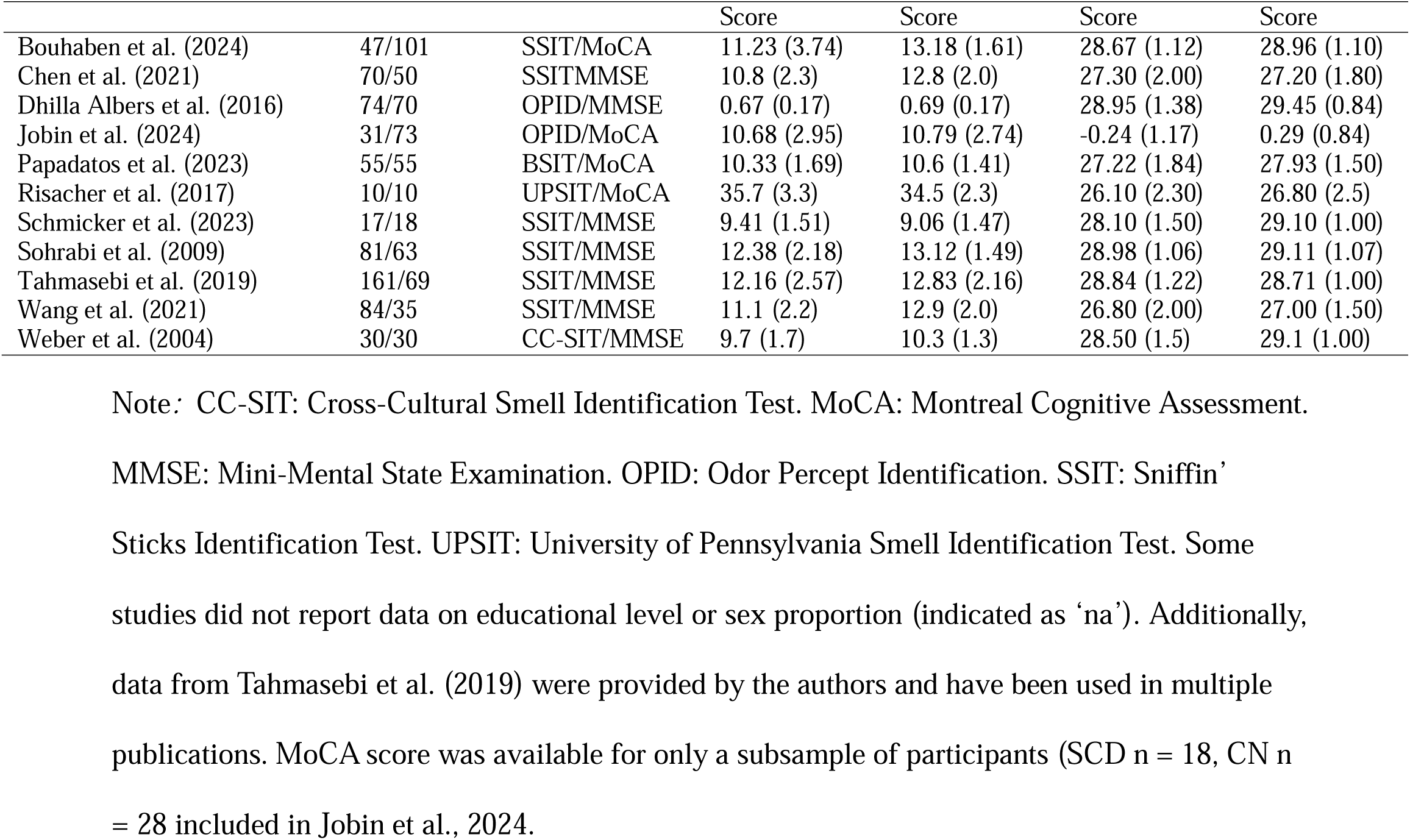
Characteristics of included studies.

### 3.2 Main Effect (Olfactory Identification)

Meta-analysis of the pooled SMD revealed that SCD groups exhibited significantly lower olfactory identification scores compared to CN groups (SMD= −0.67, 95% CI [−1.31, −0.03], *p* = .04). Variability in effect sizes across studies was observed (*I*² = 92.1%, τ² = .73) and statistically significant (Q(10) = 125.84, *p* < .001) (Figure 2).

**Figure 2:**
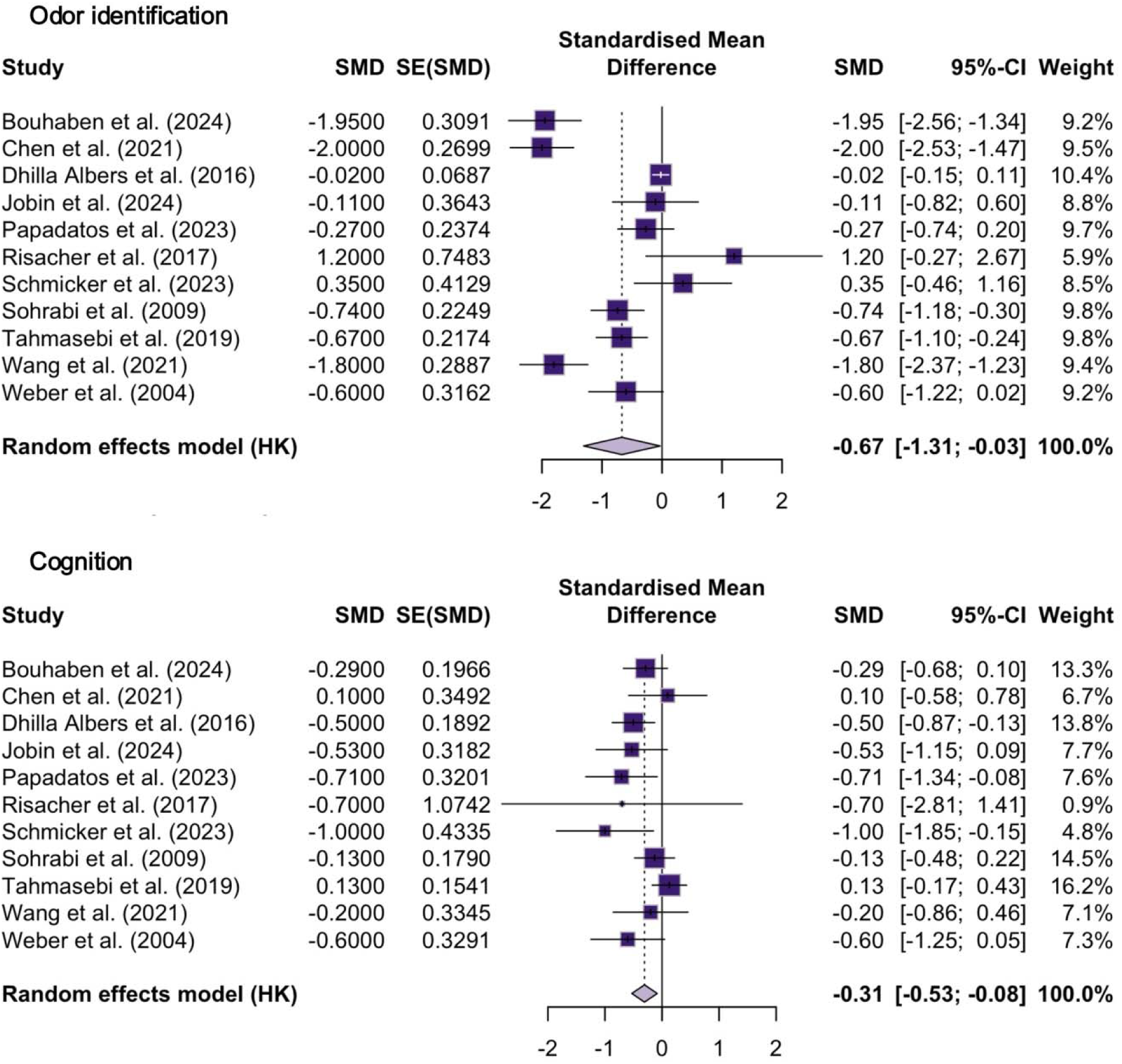
Forest plots of standardized mean differences (SMD) of odor identification and cognitive performances in individuals with SCD vs. CN participants.

Each horizontal line represents the 95% confidence interval (CI) for an individual study, with the square indicating the SMD estimate. The size of the square reflects the weight of the study in the meta-analysis. The diamond at the bottom represents the overall pooled SMD and its 95% CI, estimated using a random-effects model.

#### 3.2.1. Moderator Analysis

Meta-regression analyses were performed to identify potential sources of heterogeneity. *Age* differences between SCD and CN groups did not significantly moderate the observed differences in odor identification performance (β = 0.09, SE = 0.20, 95% CI [−0.36, 0.54], *p* = .67, *R*^2^ = 0%), suggesting that age alone does not account for the variability in effect sizes across studies (Figure 3). Neither *sex proportion (female %)* (β = −2.72, SE = 1.49, 95% CI [−6.14, 0.70], *p* = .10, *R*^2^ = 23.52%), *educational level* (years) (β = 0.03, SE = 0.39, 95% CI [−0.90, 0.95], *p* = .94, *R*^2^ = 0%), or *test modality* (*F*(4,6) = 1.56, *p* = .30) significantly moderated the differences observed in odor identification between SCD and CN groups. However, cognitive score on screening measures was a significant moderator (β = −1.79, SE = 0.64, 95% CI [−3.24, −0.34], *p* = .02, *R*^2^ = 46.47%), indicating that larger odor identification differences were observed in studies where cognitive differences between SCD and CN groups were smaller or minimal (Figure 4).

**Figure 3.**
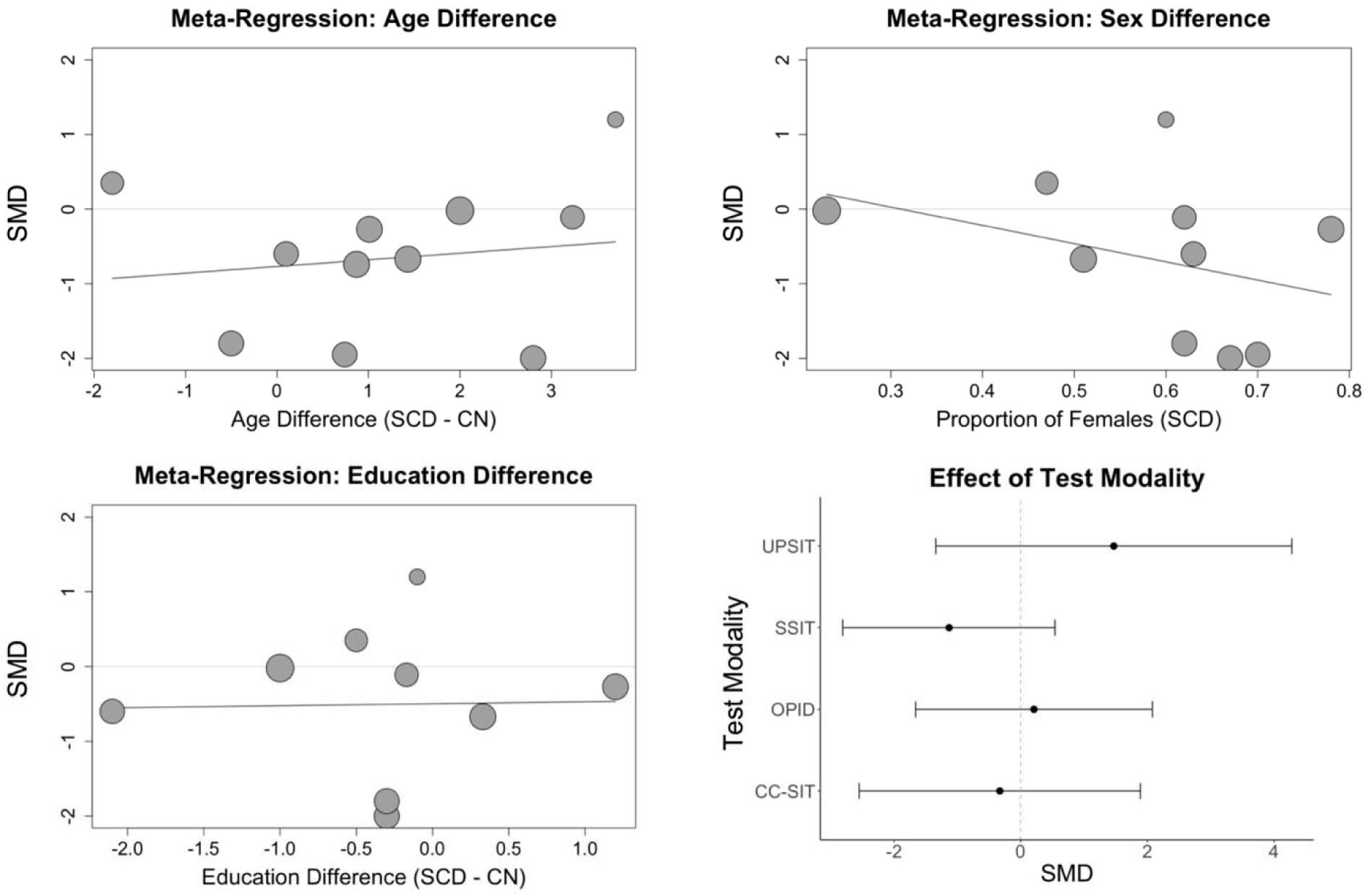
Meta-regression and Test Modality Effects on Standardized Mean Difference (SMD) in odor identification across SCD and CN groups.

**Figure 4.**
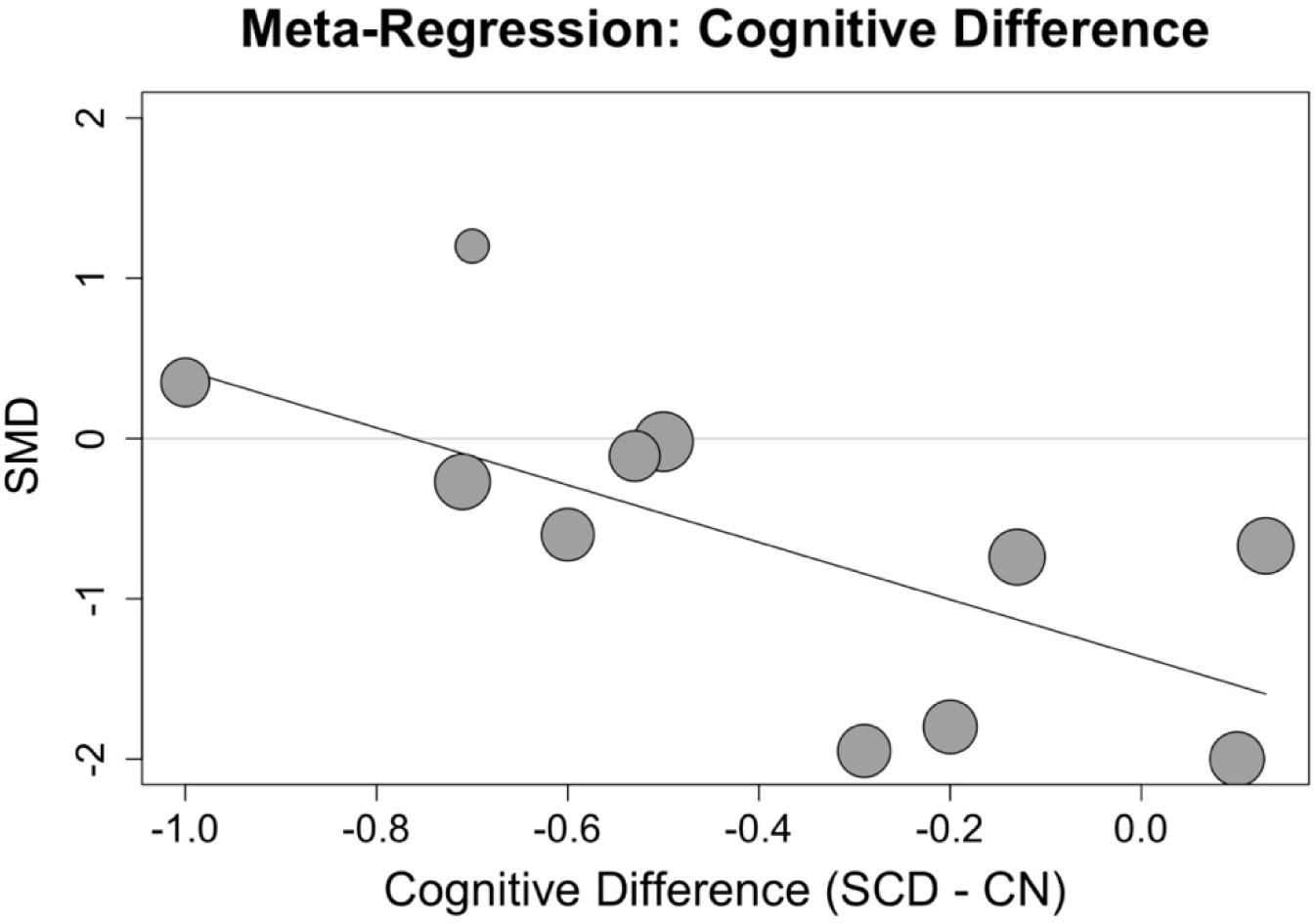
Meta-regression Effect of Cognition on Standardized Mean Difference (SMD) in odor identification across SCD and CN groups.

The four panels depict meta-regression analyses evaluating the impact of age difference (top-left), sex difference (top-right), and education difference (bottom-left). The bottom-right panel shows the effect of test modality on SMD, with different olfactory identification tests (UPSIT, SSIT, OPID, CC-SIT) represented as separate estimates with corresponding 95% confidence intervals. Negative SMD values indicate poorer olfactory performance in the SCD group compared to the CN group. Overall, none of these potential moderators reached statistical significance. Two studies did not provide information regarding sex distribution, ^49^ and one did not report educational level; ^49,50^ these studies were excluded from the respective moderator analyses.

Negative SMD values suggest poorer olfactory performance in SCD compared to CN. The meta-regression analysis identified cognitive scores on screening tests as a significant moderator (β = - 1.79, *p* = .02), indicating that larger olfactory differences were observed in studies with smaller cognitive differences between groups.

#### 3.2.2. Publication Bias

Publication bias was assessed using funnel plot inspection, Egger’s test, and the trim-and-fill method. Funnel plot symmetry indicated a low likelihood of substantial small-study effects or selective reporting, which was further supported by a non-significant result from Egger’s test (*t* = −1.79, *p* = .11). The trim-and-fill analysis imputed two additional studies, resulting in an adjusted pooled standardized mean difference of −0.30 (95% CI [−1.03, 0.44], *p* = .39), which was substantially weaker and statistically non-significant compared tothe original estimate (SMD = - 0.67, 95% CI [−1.31, −0.03], *p* = .04). This adjustment suggests that the observed effect size might have been overestimated in the presence of publication bias, and raises caution regarding the robustness of the original effect (Figure 5).

**Figure 5.**
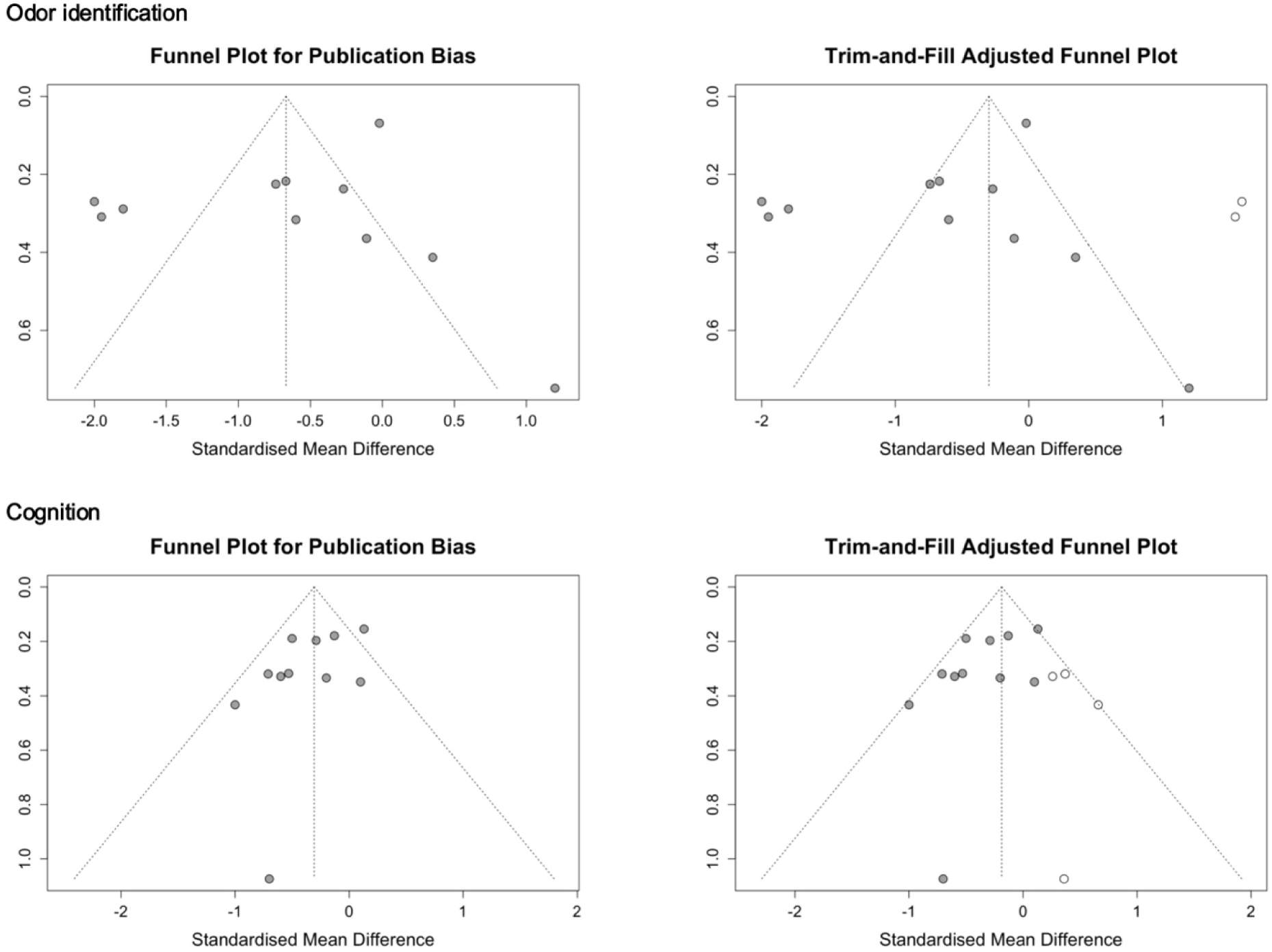
Funnel Plots Assessing Publication Bias.

For each comparison (olfactory identification and cognition), the left panel displays the funnel plot for publication bias, where each point represents an individual study’s SMD plotted against its standard error. The dotted lines indicate the expected distribution of studies in the absence of bias. Symmetry suggests minimal small-study effects or selective reporting. The right panel shows the trim-and-fill-adjusted funnel plot, with open circles representing imputed studies added to correct for potential asymmetry. The adjusted pooled effect size was smaller than the original estimate, suggesting possible overestimation due to publication bias. The wide confidence interval of the adjusted estimate includes zero, indicating that the association may not be significant after accounting for missing studies.

### 3.3. Main Effect (Cognitive Score on Screening Tests)

The meta-analysis of cognitive differences between SCD and CN also revealed a significant effect (SMD = −0.31, 95% CI [−0.53, −0.08], *p* = .01). Heterogeneity was moderate (*I*² = 39.3%, τ² = .05, *p* = .21) but not statistically significant (Q(10) = 16.48, *p* = 0.09), indicating that cognitive differences between SCD and CN are consistent across studies (Figure 2). As heterogeneity remained below the critical threshold (I² < 75%), no moderator analysis was deemed necessary. Overall, the odor identification differences (SMD = −0.67) show a larger effect size than cognitive differences (SMD = −0.31).

#### 3.3.1. Publication Bias

Funnel plot inspection showed no strong evidence of asymmetry, and Egger’s test was again non-significant (*t* = −1.88 *p* = .09), indicating a low likelihood of publication bias (Figure 4). The trim-and-fill analysis imputed four additional studies, resulting in an adjusted pooled SMD of −0.19 (95% CI [−0.42, 0.05], *p* = .11), which, although weaker and non-significant, remained in the same direction.

## 4. Discussion

Here we report that individuals with SCD exhibited lower odor identification scores compared to CN participants, extending the hypothesis that olfactory deficits may serve as an early marker of the presence of AD pathology and future cognitive decline during Stage 2 (Subjective Cognitive Decline). The effect size for group differences in odor identification (SMD = −0.67) was larger than that observed for general cognitive screening tests (SMD = −0.31), suggesting that olfactory impairment may be a more sensitive measure of subtle neurodegenerative changes in individuals with SCD. Heterogeneity was observed across studies in odor identification differences, which could not be explained by age, sex proportion, education, or test modality. Cognitive performance significantly moderated olfactory scores, however, the direction of this association indicated that larger olfactory deficits were observed in studies where cognitive differences between SCD and CN groups were smaller. This finding suggests a distinct pattern between cognitive performance on screening tests and olfactory function in SCD, supporting the potential of odor identification as a marker for neurodegenerative changes not captured by standard cognitive screening tools. Although publication bias did not appear to have strongly influenced the results, the high level of heterogeneity highlights the need for further investigation into additional contributing factors, including genetic predispositions and fluid and/or imaging biomarkers of neurodegeneration, which may account for the variability in olfactory identification deficits in SCD populations.

This meta-analysis confirmed that odor identification is reduced in SCD compared to CN participants, confirming a trend observed in an initial meta-analysis published in 2021.^23^ In the context of the AD continuum, this finding suggests that odor identification is impaired prior to the onset of cognitive deficits associated with MCI. This result aligns with studies demonstrating associations between olfactory scores and AD tau or amyloid pathology biomarkers measured in cognitively unimpaired,^17,37^ and non-demented individuals.^38,39^ A longitudinal study showed that lower olfactory identification score correlates with baseline tau accumulation in primary olfactory region and predicted accelerated progression of tau pathology in these regions ∼2.5 years after.^17^ Similarly, olfactory identification has been associated with hippocampal volume and entorhinal cortex thickness in patients with MCI^13–15^ and individuals with SCD without cognitive impairment.^16^ These findings suggest that odor identification tests could be among the earliest objective measurements of AD-related damage to brain regions involved in olfactory processing, occurring before the observable cognitive deficits.

Moderator analyses revealed that age, sex, education, and olfactory test modality did not significantly moderate the effect size. Cognitive performance on screening tests emerged as a significant moderator, although the direction of the association indicated that larger differences in odor identification performance were observed in studies where cognitive differences between SCD and CN groups were smaller. This finding reflects that olfactory identification deficits are detectable when cognitive decline is absent and that cognitive performance remains within the normal range on standard screening tests. While odor identification is not positively correlated with cognitive scores on general screening tests in this meta-analysis, prior research has shown significant positive associations between odor identification and declarative memory, which is typically the first cognitive domain affected in the course of AD.^40^ Longitudinal studies have demonstrated that lower odor identification scores predict future cognitive decline^18,41,42^ and conversion to MCI.^19,43^ Taken together, these findings support the potential role of odor identification as an early marker of neurodegenerative changes that may precede the onset of cognitive decline and MCI in at-risk individuals.

Another potential explanation for this inverse association is the differential influence of cultural bias on cognitive versus olfactory measures.^44^ While odor identification can be affected by prior sensory experience and familiarity with specific odors, cognitive screening tests might be influenced by language, educational background, cultural knowledge, and test-taking conventions. These factors may reduce the sensitivity and variability of cognitive scores across diverse populations, attenuating group differences. In contrast, olfactory tests may preserve greater discriminatory power despite cultural variation, contributing to the larger effect sizes observed in odor identification. Future studies should continue to explore how olfactory measures can be optimized for diverse populations and validated as early, culturally robust biomarkers of neurodegeneration.

It is important to note that reduced odor identification function is not specific to AD and should not be used as a stand-alone diagnostic tool. Several other conditions can impair olfaction, including neurological disorders such as Parkinson’s disease, and traumatic brain injury; as well as non-neurological conditions like chronic rhinosinusitis, among others.^45^ Therefore, interpretation of olfactory testing in the context of early AD detection must be approached with caution. Two meta-analyses demonstrated that among olfactory functions, olfactory identification is more specifically damaged in AD^9^ and MCI^10^ compared to a broader loss of smell observed in conditions affecting the peripherical olfactory system^45^ or in Parkinson’s disease.^9^ Furthermore, a recent study showed that while a higher odor identification score was associated with a higher probability of not having MCI (79%), lower score has limited sensitivity (58%) in distinguishing MCI from SCD.^46^ This suggests that the absence of an olfactory identification deficit lowers the likelihood of AD, whereas a reduced score should lead to further investigation. In such cases, more invasive and expensive diagnostic tools—such as, blood-based or CSF biomarkers, structural MRI, PET scans, comprehensive neuropsychological testing, and otorhinolaryngological examination—should be considered. Odor identification might serve as a low-cost, non-invasive first-line screening tool for brain health, and low score should trigger more in-depth evaluation to rule out other modifiable causes of olfactory dysfunction that may benefit from early intervention addressing social isolation, auditive and visual impairments, cardiovascular risk factors, depression, physical inactivity, among others.^47^

Our findings support the inclusion of an odor identification testing as an additional risk factor in the SCD+ framework, which aims to refine the identification of individuals at elevated risk of AD. This framework also includes criteria such as memory-specific complaints, onset of SCD within the past five years, age over 60, external confirmation of cognitive decline, and clinical help-seeking.^3^ Given the associations between olfactory deficits and AD biomarkers,^17,48^ medial temporal atrophy,^13–15^ and longitudinal cognitive decline,^18–20,42^ integrating odor identification into the SCD+ criteria could enhance the stratification of risk for preclinical AD. Future studies should investigate whether combining olfactory testing with the SCD+ framework improves the prediction of progression to MCI and AD dementia.

### 4.1 Limitations

The main limitation of this meta-analysis is that while meta-regression identified cognitive performance as a significant moderator, other potential contributors to heterogeneity such as genetic risk (e.g., APOE-ε4 status), neuropathological burden, or environmental factors were not assessed due to limited available data. These potential variables could have enhanced the comprehension of the significant heterogeneity observed in odor identification differences between groups (I² = 92.1%). Future research should incorporate these variables, particularly amyloid and tau biomarkers, to strengthen the specificity of olfactory impairment as a preclinical marker AD.^1^

### 4.2 Conclusion

The presence of odor identification impairments, observed alongside normal-range of cognitive performance in individuals with SCD, reinforces the hypothesis that olfactory impairment may constitute an early marker of AD preceding MCI. These findings align with previous research associating odor identification impairment with AD biomarkers and longitudinal cognitive decline. Future longitudinal research should explore whether combining odor identification testing with the SCD+ criteria improves the accuracy and precision of predicting AD pathology, future cognitive decline, and progression to MCI or AD dementia.

## Supporting information

Supplemental references

## Data Availability

All data produced in the present study are available upon reasonable request to the authors.

## Acknowledgements

The authors thank the researchers of the primary studies included in this meta-analysis for their collaboration and acknowledge the participants of those studies for their contributions.

## Consent Statement

Consent was not necessary as this study in a meta-analysis. All primary studies included in this meta-analysis obtained informed consent from their participants and reported this in their respective publications.

