## Supplemental Table 1 for "Lower Odor Identification in Subjective Cognitive Decline: A Meta-analysis"

Supplementary Table 1. Study Quality Assessment.

| Authors (Year) | Participants’ selection | Comparability  between groups | Ascertainment of  olfaction/cognition | Risk bias according  to the NOS |
| --- | --- | --- | --- | --- |
| Bouhaben et al. (2024) | 3/4 | 0/2 | 2/3 | 5/9 Moderate. |
| Chen et al. (2021) | 4/4 | 2/2 | 2/3 | 8/9 Low. |
| Dhilla Albers et al. (2016) | 3/4 | 2/2 | 2/3 | 7/9 Low. |
| Jobin et al. (2024) | 2/4 | 2/2 | 2/3 | 6/9 Moderate. |
| Papadatos et al. (2023) | 3/4 | 2/2 | 2/3 | 7/9 Low. |
| Risacher et al. (2017) | 3/4 | 2/2 | 2/3 | 7/9 Low. |
| Schmicker et al. (2023) | 3/4 | 0/2 | 2/3 | 5/9 Moderate. |
| Soharabi et al. (2009) | 3/4 | 2/2 | 2/3 | 7/9 Low. |
| Tahmasebi, et al. (2019) | 3/4 | 0/2 | 2/3 | 5/9 Moderate. |
| Wang et al. (2021) | 4/4 | 2/2 | 2/3 | 8/9 Low. |
| Weber (2003) | 3/4 | 2/2 | 2/3 | 7/9 Low. |

Supplementary Table 1 summarizes the quality assessment of each study based on key criteria: (1) Participants’ selection, which evaluates recruitment methods, use of neuropsychological assessments, and health history documentation; (2) Comparability between groups, assessing whether studies controlled for key demographic and clinical factors such as age, sex, and education; (3) Ascertainment of olfactory identification and cognition, examining the reliability and consistency of assessment methods across study groups; and (4) Overall risk of bias, categorized as low, moderate, or high based on these criteria.
